# Immortal time bias reproduces the reported survival benefit of conversion surgery in stage IV gastric cancer: a simulation study

**DOI:** 10.64898/2026.09.01.26361986

**Authors:** Birendra Kumar Sah, Chen Li, Jian Li, Zhenggang Zhu

## Abstract

**Background:** Conversion surgery for stage IV gastric cancer is supported by a pooled overall survival hazard ratio of 0.36 (95% confidence interval 0.32–0.40) and, in the largest international cohort, median survival of 36.7 versus 12.5–13.8 months on chemotherapy. Survival is measured from diagnosis; the median diagnosis-to-gastrectomy interval is 124 days, which patients must survive to be counted surgical.

**Methods:** We simulated cohorts of 3,177 stage IV gastric cancer patients from published parameters: background median survival 14.5 months; median diagnosis-to-surgery interval 124 days (category-specific 92–174 days). Surgery had no effect (true hazard ratio 1.00 by construction). Data were analysed as the literature analyses them (exposure fixed at baseline, follow-up from diagnosis), and by time-varying Cox and landmark analysis. Confounding by indication was added in a second scenario.

**Results:** Under immortal time bias alone the naive analysis returned a hazard ratio of 0.794 (95% simulation interval 0.743–0.851), median survival 16.8 versus 12.8 months. Time-varying Cox recovered 1.000 and landmark analysis 1.000–1.004. Bias scaled with the interval: 0.849 at 92 days, 0.715 at 174 days. Adding confounding, the naive estimate fell to 0.601 (0.560–0.644) at strength 0.5 and 0.356 (0.323–0.385) at strength 1.5, overlapping the published estimate; median survival 21.9 versus 8.7 months. Correcting immortal time alone left residual bias (hazard ratio 0.439).

**Conclusions:** The reported survival advantage of conversion surgery is reproducible where the operation does nothing; published estimates cannot distinguish benefit from bias. Resolving this requires individual patient data analysed with methods that assign person-time correctly, or completion of JCOG2301.

## Introduction

Conversion surgery (resection with curative intent after systemic therapy in initially unresectable stage IV gastric adenocarcinoma) has moved from anecdote toward standard practice within a decade.

The most recent systematic review identified 36 studies comprising 3,177 patients and reported a pooled overall survival hazard ratio of 0.36 (95% CI 0.32–0.40), I² = 27%, across 32 studies [1]. Median overall survival ranged from 14.4 to 60.0 months in surgical groups and 4.7 to 19.9 months in non-surgical groups. A second synthesis of 12 cohorts reported advantages of similar magnitude [2]. Earlier syntheses, including one confined to peritoneal metastasis, reached the same conclusion [3, 4]. International expert consensus recorded 88.3% agreement that conversion therapy confers survival benefit in selected responders achieving R0 resection [5]. The only randomised trial of gastrectomy in incurable gastric cancer, REGATTA, was closed at interim analysis for futility [6].

The largest dataset, CONVO-GC-1, registered 1,902 patients across 55 institutions in Japan, Korea and China and analysed 1,206, reporting median survival of 36.7 months (95% CI 34.4–40.0), rising to 56.6 months after R0 resection [7].

Three features of that study establish the problem addressed here, and all three are stated within it.

First, survival is measured from diagnosis, defined as time to death, irrespective of cause, from the date of diagnosis [7]. Second, the interval to surgery is substantial: the reported median duration of preoperative chemotherapy, explicitly the period between disease diagnosis and gastrectomy, was 124 days (IQR 81–209), and 92, 135.5, 158 and 174 days across Yoshida categories 1–4 [7], a biological classification proposed in this journal for precisely this population [8]. Third, the cohort is defined by having reached surgery: of 1,902 registered patients, 72 were excluded for incomplete data and 624 because they underwent surgery without preoperative chemotherapy, leaving 1,206 who received chemotherapy and then an operation. All survived to surgery. There is no comparator arm. Follow-up therefore begins at diagnosis while exposure is determined by an event occurring a median of four months later. Death in that interval is impossible for anyone classified as surgical, since such a patient never reaches the operating table. This person-time is immortal, and attributing it to the surgical group inflates apparent survival irrespective of treatment effect [9]. The canonical demonstration (an apparent mortality benefit of statins that disappeared once the immortal interval was reassigned) established the magnitude such misalignment can generate [10].

The comparison this invites is made within CONVO-GC-1 itself: its Introduction cites median survival of 12.5–13.8 months in phase III trials of first-line chemotherapy for M1 disease, described as unsatisfactory; its Results report 36.7 months [7]. The first describes all patients diagnosed; the second describes those who survived four months of chemotherapy and were fit for major surgery.

Existing syntheses acknowledge bias but name a different mechanism. Chan et al. [1] cite heterogeneous selection criteria for conversion surgery that may favour patients with good tumour biology; Wu et al. [2] concede that their findings may reflect inherent prognostic differences between groups, since surgery was feasible only for chemotherapy responders. Both describe confounding by indication. Neither names immortal time; neither corrects for it.

Immortal time bias has been formally addressed in conversion surgery for pancreatic ductal adenocarcinoma, where propensity score matching, time-dependent covariate Cox regression and landmark analysis were applied together [11], and in biliary tract cancer, where operative status was modelled as a time-varying covariate [12]. In stage IV breast cancer, early observational studies suggested a survival benefit from primary tumour resection that did not survive correction [13], and a subsequent simulation study in which the true relationship between resection and mortality was specified quantified the magnitude of the bias directly [14]. In stage IV colon cancer, a simulation comparing time-fixed exposure against time-varying exposure, delayed entry and landmark methods, applied to a population-based cohort under a target trial emulation framework, found the same pattern and showed that the magnitude of the bias scales with the underlying risk of early death [15]. We are aware of no equivalent analysis in gastric conversion surgery.

We therefore asked: in a world where conversion surgery has no effect on survival, what hazard ratio would the published methods report?

## Methods

Reported according to the ADEMP framework for simulation studies [16].

### Aims

To quantify the hazard ratio and median survival difference generated by immortal time bias, alone and combined with confounding by indication, under a specified null treatment effect.

### Data-generating mechanism

Cohorts of n = 3,177 (matching the largest meta-analysis [1]) were simulated from date of diagnosis. Latent survival time T followed a Weibull distribution with scale calibrated to a median of 14.5 months, slightly above the 12.5–13.8 months reported across recent phase III trials of first-line chemotherapy for M1 disease [7]. This choice is conservative with respect to our own argument: a shorter background median would lengthen the immortal interval relative to survival and generate more bias, not less. The shape parameter, 1.30 in the primary analysis, is not directly reported in this population; we therefore checked the assumed value against published chemotherapy-alone survival. A two-parameter Weibull distribution was fitted to two published survival points from the same CheckMate 649 chemotherapy-alone subgroup (PD-L1 combined positive score ≥ 5, for which this arm’s median of 11.1 months is stable across successive follow-up analyses). Using 19% alive at 24 months and 10% alive at 36 months as the two calibration points yields shapes of 1.13 and 1.02 respectively, a fitted range with midpoint 1.08 [17]. All analyses were repeated across shapes of 1.02–1.30. Survival time was drawn independently of surgical status: surgery had no effect, and the true hazard ratio was 1.00 by construction.

Scheduled surgery time C followed a log-normal distribution with median 4.07 months (124 days; σ = 0.35 on the log scale), taken from the diagnosis-to-gastrectomy interval reported in CONVO-GC-1 [7]. Sensitivity analyses used category-specific values (92, 135.5, 158, 174 days) and peritoneal-specific values (84 days for P0CY1; 167 days for directly visualised dissemination).

Patients were independently designated as offered surgery with probability p; surgery occurred only if offered and T > C. Administrative censoring was applied at 60 months. Under Scenario A, the probability of being offered surgery was independent of prognosis, isolating immortal time bias. Under Scenario B, the log-odds of being offered surgery increased linearly with latent survival standardised on the log scale, at strengths 0.5–2.0, adding confounding by indication. Because the induced bias depends on the behaviour of the selection function in the upper tail of the survival distribution, the choice of scale is consequential and is reported here rather than left implicit: under otherwise identical parameters at strength 1.5, standardising raw survival time yields a naive hazard ratio of 0.27 and rank-standardisation yields 0.29, against 0.36 on the log scale.

### Estimand

The marginal hazard ratio for surgery versus no surgery, and the difference in median overall survival between groups.

### Methods evaluated

Naive Cox regression: surgical status as a fixed baseline covariate with follow-up from diagnosis, reproducing the published approach and the time-zero stated in CONVO-GC-1. Time-varying Cox regression: person-time before surgery assigned to the unexposed group. Landmark analysis [18–20]: at 3.02, 4.07 and 5.72 months, the empirically anchored intervals from CONVO-GC-1, excluding patients dying before the landmark.

### Performance measures

Median and 2.5th–97.5th percentile of estimates across replicates (“simulation intervals”), and Kaplan–Meier median survival by group. Monte Carlo standard errors for each median estimate were obtained by bootstrap resampling of the replicate distribution (4,000 resamples); they were ≤ 0.004 throughout and 0.0007 for the headline naive hazard ratio at confounding strength 1.5, confirming that 500 replicates resolve the reported differences far more finely than the effects being described. The primary scenarios reported in Tables 1–3 and Figs. 1–3 each used 500 replicates. The supporting sensitivity analyses used fewer, as they are reported only as ranges or contrasts: 400 replicates for the hazard-shape analysis, 300 for the landmark decomposition, 200 and 150 for the response-mediated models, and 60 for the standardisation-scale diagnostic. Analyses used Python 3. Because every model involves a single binary covariate, the naive, time-varying and landmark Cox models were fitted with a dedicated Newton–Raphson solver for the Breslow partial likelihood rather than by generic software; this is the same likelihood and agreed with lifelines 0.30.3 to within 1.4 × 10 across all model types and both confounding settings. Kaplan–Meier medians used lifelines 0.30.3. Code and replicate-level output are available from the corresponding author on reasonable request.

**Fig. 1.**
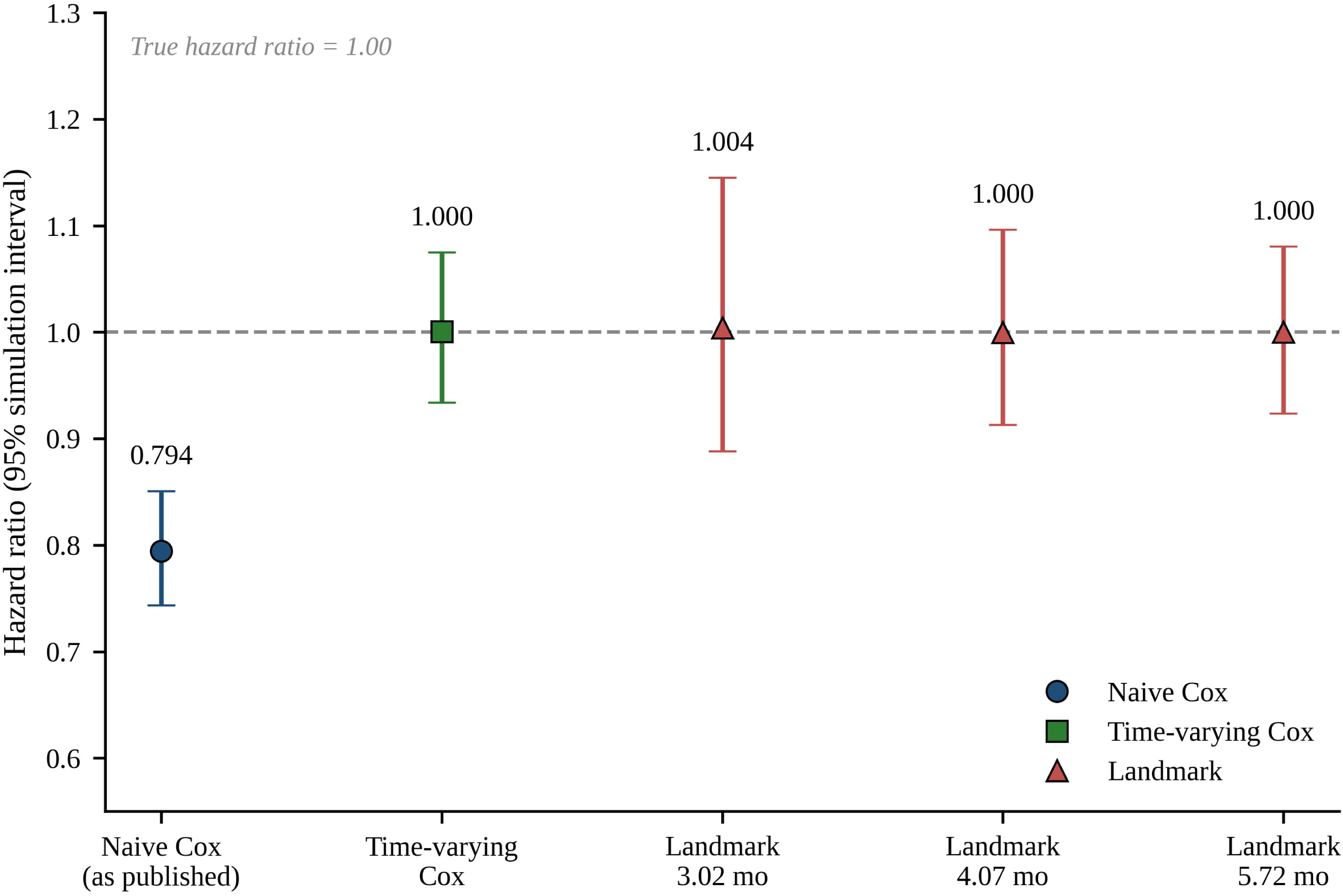
Hazard ratios recovered under pure immortal time bias by analytical method. Points show medians across 500 replicates; bars show 2.5th–97.5th percentiles. The dashed line marks the true hazard ratio of 1.00. Surgery was specified to have no effect on survival

**Fig. 2.**
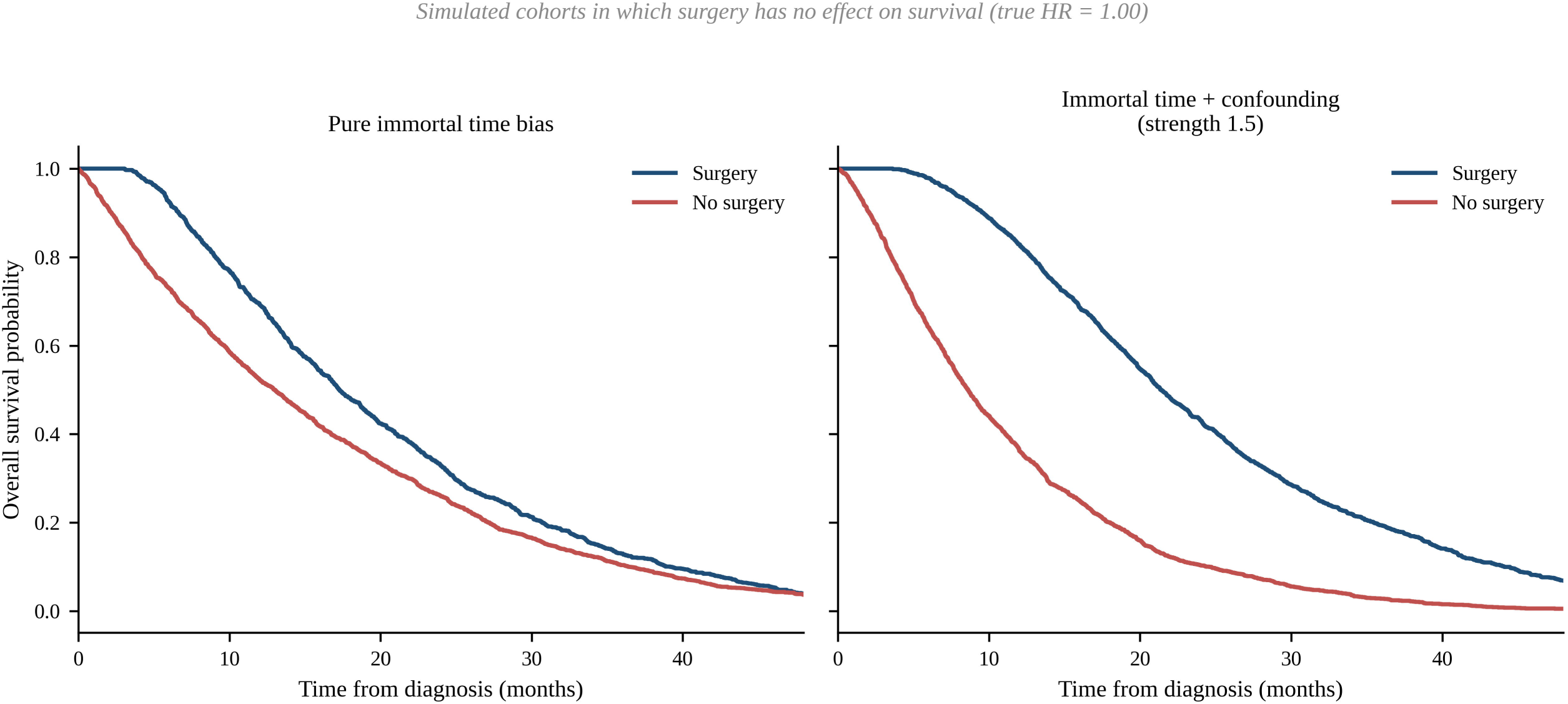
Kaplan–Meier curves from simulated cohorts in which surgery has no effect on survival. a Pure immortal time bias. b Immortal time bias with confounding by indication (strength 1.5)

**Fig. 3.**
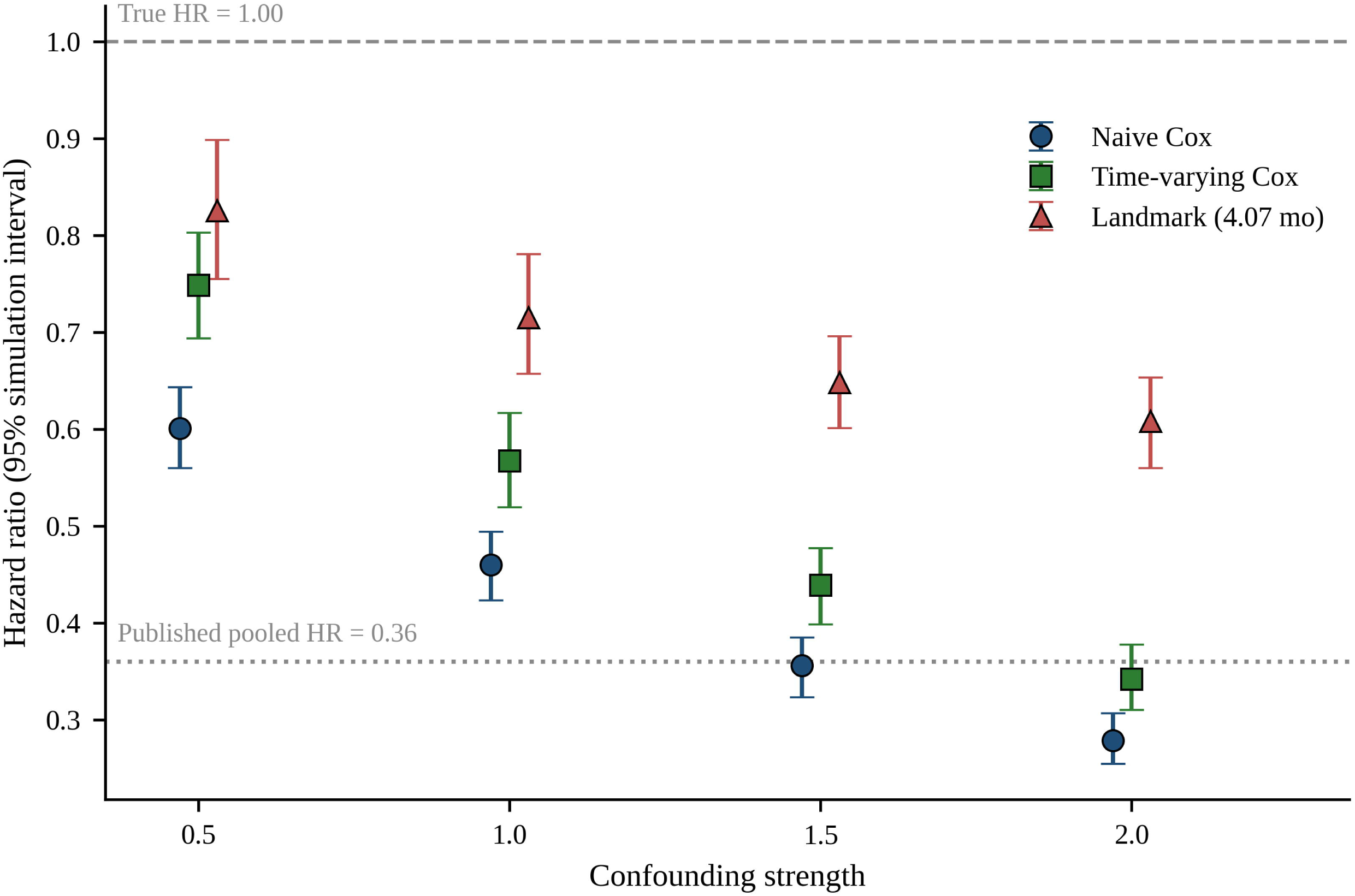
Estimated hazard ratio against strength of confounding by indication for three analytical approaches. The dashed line marks the true hazard ratio of 1.00; the dotted line marks the published pooled estimate of 0.36

**Table 1.** Hazard ratios and survival under pure immortal time bias (Scenario A; true hazard ratio = 1.00)

| **Analysis** | **HR** | **95% SI** |
| --- | --- | --- |
| Naive Cox (as published) | 0.794 | 0.743–0.851 |
| Time-varying Cox | 1.000 | 0.934–1.075 |
| Landmark, 3.02 mo | 1.004 | 0.888–1.145 |
| Landmark, 4.07 mo | 1.000 | 0.913–1.096 |
| Landmark, 5.72 mo | 1.000 | 0.924–1.081 |
| **Median OS, surgery group (mo)** | **16.8** | **16.0–17.6** |
| **Median OS, no-surgery group (mo)** | **12.8** | **12.1–13.6** |
| **Median OS, whole cohort (mo)** | **14.5** | **14.0–15.1** |
| **Conversion rate** | **0.39** | **0.37–0.41** |

HR hazard ratio, OS overall survival, SI simulation interval
Five hundred replicates of $n = 3,177$ . Monte Carlo standard error of each median hazard ratio $\leq 0.004$

**Table 2.** Naive hazard ratio by diagnosis-to-surgery interval (true hazard ratio = 1.00)

| **Cohort** | **Interval (days)** | **Months** | **Naive HR** | **Median OS, surgery vs no surgery (mo)** |
| --- | --- | --- | --- | --- |
| P0CY1 (peritoneal) | 84 | 2.76 | 0.861 | 15.9 vs 13.4 |
| Yoshida category 1 | 92 | 3.02 | 0.849 | 16.0 vs 13.3 |
| **All categories** | **124** | **4.07** | **0.794** | **16.8 vs 12.7** |
| Yoshida category 2 | 135.5 | 4.45 | 0.774 | 17.0 vs 12.6 |
| Yoshida category 3 | 158 | 5.19 | 0.741 | 17.6 vs 12.3 |
| P(+)-direct (peritoneal) | 167 | 5.49 | 0.729 | 17.8 vs 12.1 |
| Yoshida category 4 | 174 | 5.72 | 0.715 | 18.0 vs 12.0 |

HR hazard ratio, OS overall survival
Intervals taken from CONVO-GC-1 [7]. Rows ordered by interval length. Five hundred replicates per value.
Monte Carlo standard error of each median hazard ratio $\leq 0.002$

**Table 3.**
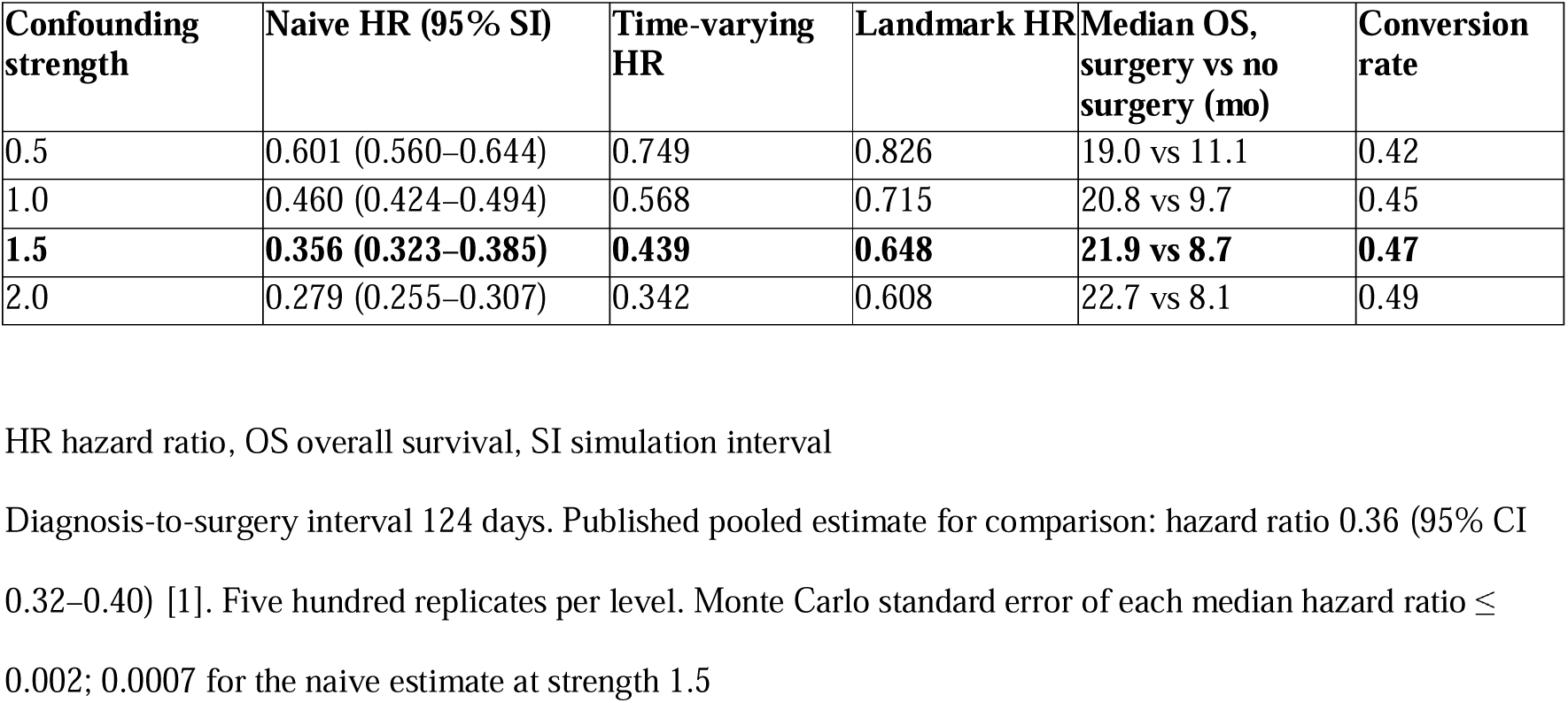
Immortal time bias with confounding by indication (Scenario B; true hazard ratio = 1.00)

## Results

### Immortal time bias alone

Under pure immortal time bias, the naive analysis returned a hazard ratio of 0.794 (95% simulation interval 0.743–0.851), an apparent 20.6% reduction in the hazard of death from an intervention with no effect (Table 1, Fig. 1). Median overall survival was 16.8 months in the surgical group versus 12.8 months in the non-surgical group, against a whole-cohort median of 14.5 months, with a conversion rate of 39%. Time-varying Cox regression recovered a hazard ratio of 1.000 and landmark analysis 1.000–1.004.

Simulated survival curves from cohorts in which surgery had no effect are shown in Fig. 2.

### Bias scales with the diagnosis-to-surgery interval

Applying the category-specific intervals from CONVO-GC-1, the naive hazard ratio fell monotonically from 0.849 at 92 days to 0.715 at 174 days (Table 2). Peritoneal-specific values gave 0.861 for P0CY1 disease (84 days) and 0.729 for directly visualised dissemination (167 days). Bias was greatest in categories 3 and 4 and in macroscopic peritoneal disease, the settings in which conversion surgery is most contested.

The same gradient held across the wider parameter space, with naive hazard ratios between 0.596 and 0.887 depending on interval and conversion rate.

Bias magnitude depended on the assumed hazard shape, but not in a direction favourable to our argument. Across the fitted range and the assumed value (shapes of 1.02, 1.08, 1.13 and 1.30), pure immortal time bias produced naive hazard ratios of 0.73, 0.74, 0.75 and 0.79 respectively, while the confounded estimate at strength 1.5 varied only between 0.34 and 0.35. The assumed shape is therefore the conservative choice: at every fitted value, immortal time alone generates more apparent benefit, not less, and the reproduction of the published estimate is insensitive to this parameter.

### Immortal time plus confounding by indication

Adding confounding by indication, the naive estimate fell to 0.601 (0.560–0.644) at strength 0.5 and to 0.356 (0.323–0.385) at strength 1.5, overlapping the published pooled estimate of 0.36 (95% CI 0.32–0.40) (Table 3, Fig. 3). Median survival at that strength was 21.9 versus 8.7 months, within the published ranges of 14.4–60.0 and 4.7–19.9 months.

Correcting immortal time alone was insufficient: at strength 1.5 the time-varying estimate remained 0.439. Landmark analysis returned an estimate closer to the null (0.648), but this reflects exposure misclassification rather than confounding control. At a landmark set to the median diagnosis-to-gastrectomy interval, approximately half of all patients who ultimately underwent conversion surgery had not yet been operated and were classified as unexposed, diluting the exposure contrast. When patients operated after the landmark were excluded, landmark and time-varying estimates converged (0.44 versus 0.44), as expected given that neither method addresses confounding. The residual difference tracked the proportion of not-yet-operated patients across landmark times from 2.00 to 8.00 months. Even a landmark set near the conventional median time to surgery retains substantial misclassification bias (shift 0.212 at 4.07 months); earlier landmarks are more vulnerable still.

### Single-arm comparison against a historical median

Comparing the operated cohort’s median survival against the whole-population median (the structure by which 36.7 months is read against 12.5–13.8 months) produced apparent gains of 2.3 months under pure immortal time, rising to 4.5, 6.2 and 7.4 months at confounding strengths 0.5, 1.0 and 1.5, from zero true effect.

## Discussion

In a simulated world where conversion surgery does nothing, the approach used throughout the published literature reports a hazard ratio of 0.794 from immortal time alone, and 0.356 (overlapping the published estimate) once moderate confounding is added. The accompanying medians fall within the ranges reported across 36 real studies.

We emphasise what this does and does not show. It does not show that conversion surgery is ineffective; complete resection of residual disease in a chemosensitive patient is biologically coherent and may confer real benefit. It shows that the published evidence cannot distinguish a large benefit from no benefit, because a design that generates a hazard ratio of 0.36 under the null cannot estimate an effect near 0.36.

### A comparison where benefit is implausible

The clearest illustration is not the headline figure. Chan et al. report that patients undergoing non-R0 resection (where disease was left behind) still showed superior survival against no conversion surgery: hazard ratio 0.67 (95% CI 0.53–0.86), 8 studies, 717 patients [1]. The authors attribute this to the benefits of tumour debulking or to inherently favourable tumour biology.

There is no coherent mechanism by which an incomplete resection reduces mortality by a third. Our simulation supplies the alternative: pure immortal time produced 0.794 and mild confounding 0.601. The observed 0.67 lies between them. This is not a marginal subgroup: 26.6% of conversion surgery patients underwent non-R0 resection [1].

### Low heterogeneity as a signature

Chan et al. report I² = 27% across 32 studies, with meta-regression showing no association of the pooled effect with study design (P = 1.00), sample size (P = 0.13) or publication year (P = 0.67) [1]. These are presented as robustness.

An alternative reading is available. Studies with genuinely different treatment effects (different regimens, eras, case mixes, techniques) should not agree that closely. Studies sharing an identical structural feature should. The effect is invariant to everything tested, and the one thing untested is the design feature common to all.

### Why appraisal did not detect it

Both meta-analyses used the Newcastle–Ottawa Scale [1, 2, 21]. The convention extends beyond gastric cancer; a 2026 meta-analysis of conversion surgery in oesophageal cancer used it likewise [22].

The Newcastle–Ottawa Scale contains no domain assessing alignment between start of follow-up and start of intervention. Its closest item, demonstration that the outcome of interest was not present at the start of the study, addresses prevalent outcomes, not exposure-time misalignment. In the quality table of Wu et al., all twelve included studies scored maximum on that item, with totals of 7–8 of 9 [2]. Studies embedding immortal time by design were formally rated high quality.

ROBINS-I includes bias in selection of participants into the study as an explicit domain covering this misalignment [23]. The choice of instrument determines whether the dominant threat to validity is visible. This generalises to delayed metastasectomy, transplantation after bridging therapy, and salvage resection after chemoradiation.

### Limitations

Our data-generating model is a simplification, omitting competing risks, treatment switching and heterogeneity not captured by a single latent survival variable. Confounding strength is unobservable; the value of 1.5 reproducing the published estimate is an existence proof that plausible bias suffices, not a measurement of bias present.

Selection to surgery in our model operates on latent survival directly, whereas clinically it operates through response to chemotherapy, correlated with prognosis but distinct from it. We therefore repeated the analysis with selection acting on a response variable correlated with the underlying prognostic index. Two findings bear on interpretation. The immortal-time component was mechanism-invariant: wherever the naive estimate equalled 0.36, time-varying regression returned 0.44 regardless of how confounding was generated. The existence proof, however, is mechanism-dependent. When response and prognosis were only weakly correlated, even near-deterministic selection on response could not drive the naive estimate below 0.48. Reproducing 0.36 therefore requires chemotherapy response to be a reasonably strong prognostic marker in this population: defensible for metastatic gastric cancer, but an assumption rather than a demonstration. The simulation cannot establish that no true effect exists; it can only show that published designs could not detect the difference. Finally, we did not search non-English databases when assessing whether this bias has been addressed elsewhere; we note that Chan et al. likewise excluded non-English studies while assembling the field’s definitive synthesis [1].

### Implications

Guideline and consensus authors, including those of the Japanese gastric cancer treatment guidelines [24], should report the pooled hazard ratio and the 36.7-month median as upper bounds, explicitly uncorrected for immortal time, and cease juxtaposing single-arm surgical survival against unselected historical medians; CONVO-GC-1 was designed with operative complications as its primary endpoint, and its survival figure has acquired a weight its design did not intend to bear [7].

The CONVO-GC consortium holds 1,206 patients with diagnosis and surgery dates recorded. A target trial emulation of those data [25, 26] would resolve in one analysis what no further aggregate-data synthesis can address; numbers at risk are printed beneath every published curve, so external reconstruction is also feasible [27].

Future primary studies should pre-specify handling of immortal time, report the diagnosis-to-surgery interval, define the comparator as responding patients not operated rather than all non-operated patients, and analyse by intention to resect rather than conditionally on R0 achievement. Systematic reviewers should use ROBINS-I wherever an intervention follows a variable interval of prior therapy.

JCOG2301 is now recruiting, comparing chemotherapy alone against conversion surgery after remarkable response [28], the design Wu et al. independently called for [2]. It is the right trial, but will take years to report, and in the interim the field treats a pooled hazard ratio of 0.36 as a provisional answer. Our purpose is to quantify why it cannot serve as one. A caution on generalisability: 35 of the 36 studies in the largest meta-analysis were conducted in East or Southeast Asia [1].

## Conclusion

A hazard ratio of 0.36 for conversion surgery in stage IV gastric cancer is reproducible in a world where the operation has no effect. Whether conversion surgery benefits patients remains open, and answering it requires individual patient data analysed with methods that assign person-time correctly, or completion of JCOG2301. Until then, the difference between a recommendation and a hypothesis should be stated plainly.

## Data Availability

All data produced in the present study are available upon reasonable request to the authors. No patient-level data were used; all analysed data were simulated, and the code that generates them is available on request.

## Acknowledgements

We thank Clinical Research Center, Ruijin Hospital, Shanghai Jiao Tong University School of Medicine for specifying and validating the data-generating mechanism and performance measures, and for statistical review of the manuscript.

This research received no specific grant from any funding agency in the public, commercial, or not-for-profit sectors.

## Author contributions

BKS conceived the study, designed the simulation, wrote the analysis code, and drafted the manuscript. CL, ZZ contributed to the conception and framing of the study, to the interpretation of the simulated scenarios against surgical decision-making in practice, and critically revised the manuscript for clinical content. JL specified and validated the data-generating mechanism and performance measures, and provided statistical review of the manuscript. All authors critically revised the manuscript, approved the final version, and accept accountability for the work.

## Data availability

This study generated no patient-level data; all data analysed were simulated. The simulation and analysis code, and the replicate-level output supporting Tables 1–3 and Figs. 1–3, are available from the corresponding author on reasonable request.

## Ethical approval

This article does not contain any studies with human or animal subjects performed by any of the authors.

